# Identification of Cerebrospinal Fluid Proteomic Signatures Associated with Intramedullary Surgery Outcome

**DOI:** 10.1101/2023.06.23.23290571

**Authors:** Weihao Di, Wentao Gu, Xike Peng, Jiajun Shou, Yi Zhang, Shufei Ge, Jian Zhao, Shixin Gu, the Neurospine Group

## Abstract

**Importance:** Neurological function recovery after intramedullary microsurgery is susceptible to various factors. Investigating alterations in cerebrospinal fluid (CSF) protein profiles associated with spinal function recovery could provide valuable insights into prognosis and the development of postoperative therapeutic approaches.

**Objective:** The aim of this research was to identify CSF protein sets linked to recovery outcomes of intramedullary microsurgery.

**Design:** This research is an observational cohort investigation that collected data from August 2020 to March 2023.

**Setting:** The Neurospine group conducted this single-centered cohort research (http://www.chictr.org.cn, registration ID: ChiCTR2300072704). Participants were followed up for 6 months or more. CSF samples were collected pre-and postoperatively. Perioperative treatments, neurological deficits, and rehabilitation therapies were recorded.

**Participants:** The Neurospine group screened adult patients with intramedullary lesions scheduled for surgical intervention. The exclusion criteria included prior spinal surgery, neurological disorders, surgical complications, or relevant medications.

**Exposures:** The patients underwent standard intramedullary microsurgical procedures, received medications, and underwent assessments. The CSF protein profiles were examined using tandem mass spectrometry.

**Main outcomes and measures:** The evaluation of spinal function recovery involved various assessments and correlation analysis was performed. In addition, CSF protein features were identified and their clinical associations were assessed using least absolute shrinkage and selection operator (LASSO) regression modeling.

**Results:** Of the 43 patients who completed follow-up assessments, 23 were women and 20 were men, with a mean age of 42.8±11.8 years. Histological diagnoses confirmed invasive lesions in 20 patients. Twenty-three patients demonstrated stable or improved spinal function, while 20 patients experienced deterioration during the 3rd and 6th month follow-ups. As indicated by the changes in EMS scales (*P* = 0.02) and JOA scores (*P* = 0.03), functional prognosis was associated with pathological characteristics. We developed four models using the CSF proteomics datasets of noninvasive and invasive patients. These LASSO regression models exhibited high accuracy in predicting the recovery rates of the corresponding groups (*P* > 0.99).

**Conclusions and relevance:** This research demonstrates that lesion sizes and pathological characteristics are significant factors influencing spinal function recovery. The LASSO regression models developed using identified CSF protein variables for invasive and noninvasive patients, respectively, offer novel methods for the prediction of prognosis and the development of therapeutic approaches.

## Introduction

Intramedullary lesions, with tumors being a significant portion, are uncommon clinical conditions that result in spinal neurological dysfunction. In the United States, the reported incidence of intramedullary spinal cord tumors ranges from 0.31 to 0.35 per 100,000 individuals.^1, 2^ Surgical interventions typically involve making incisions in the spinal cord and performing microsurgical removal of the tumor from the spinal cord tissue.^3–6^ The surgery may also lead to impairment of spinal cord function due to the delicate nature of the spinal cord tissue. The extent of this impairment is influenced by various factors, such as the preoperative neurological status of the patient, lesion size, degree of adhesion to the spinal cord, and the operating experience of the surgeon. Currently, postoperative treatments for spinal cord injuries primarily consist of symptom-alleviating medications and rehabilitation.

On the other hand, surgical resection of intramedullary lesions presents a distinct opportunity to obtain cerebrospinal fluid (CSF) samples before and after the procedure. The analysis of these samples provides considerable potential in understanding the physiological alterations that occur after surgery and may reveal potential biomarkers for prognosis,^7, 8^ offering not only reference markers but also therapeutic targets for postoperative treatments.^9^

Isotopic chemical tags, which are tandem mass tags (TMTs), are employed to label peptides from various samples.^10, 11^ This labeling process enables the simultaneous analysis of multiple samples using mass spectrometry (MS). Tandem Mass Tag Mass Spectrometry (TMT-MS) provides a robust system for both qualitative and quantitative comparisons of protein profiles from different patients or multiple time points within a single patient.^12, 13^

Nevertheless, the abundance of proteins in CSF presents a significant technical challenge for downstream data analysis and interpretation.^14, 15^ In high-dimensional data analysis, the least absolute shrinkage and selection operator (LASSO) regression algorithm is widely employed as a feature selection method.^16–18^ It offers a distinct advantage in mitigating the risk of over-fitting and reducing model variance through simultaneous feature selection and regularization.^19^ LASSO proves particularly valuable when dealing with problems that involve numerous features (the extensive proteomics data per sample) but a relatively small sample size (the limited number of participants that meet the recruitment criteria for a cohort study).^20^

In this cohort study, we assessed the neurological recoveries of patients who underwent intramedullary surgery, monitored changes in CSF protein profiles in these patients using TMT-MS analysis, and employed LASSO penalized regression modeling for identification of CSF protein sets associated with spinal function recovery in the corresponding patient populations.

## Methods

### 1. Study design and cohort participants

This prospective cohort study was approved by the institutional ethics committee, registed (http://www.chictr.org.cn, registration ID: ChiCTR2300072704) and conducted by the Neurospine group at the Department of Neurosurgery. Before enrollment, informed consent was obtained from all participants.

From August 2020 to September 2022, the Neurospine group conducted a screening of adult patients (18 years or older) with intramedullary lesions scheduled for surgical intervention. Patients with surgical complications, prior spinal surgeries, pregnancy, neurological disorders, or medications that could impact CSF protein profiles were excluded. All participants underwent a comprehensive neurological examination, and a spinal magnetic resonance (MR) imaging was conducted to assess the extent of the lesions and confirm the diagnosis. After administering general anesthesia, CSF samples were collected via a lumbar puncture. Standard intramedullary microsurgery was conducted. This surgery involved a spinal cord incision to expose the lesion and microsurgical manipulation to detach and remove the tumor. To minimize spinal cord strain and damage during tumor separation, we exposed the spinal cord in the posterior median sulcus, where nerve conduction tracts are weak. Before suturing the dura, a second CSF sample from the same patient was obtained from the subarachnoid area. Postoperatively, patients were assessed daily for any neurological deficits and complications. Subsequently, the patients were followed up for 6 months and their neurological function were evaluated. An MR scan was conducted at the 6th month follow-up. Additionally, all patients were also subjected to long-term follow-ups every three months, and comprehensive neurological examinations were scheduled every six months during clinical visits for all patients.

### 2. Demographic and clinical data

We collected demographic information and clinical data of all participants, including sex, age, symptoms, pathology results, degree of lesion excision, adjuvant, perioperative medication, surgery date, and rehabilitation therapy. Multiple assessments, such as the American Spinal Injury Association (ASIA) Impairment Scale (AIS),^21^ Neurological Level of Injury (NLI),^21^ Neck Disability Index (NDI),^22^ European Myelopathy Score (EMS),^23^ and Japanese Orthopaedic Association (JOA) scores,^24–26^ were conducted. Furthermore, the functional recovery of the patients was calculated and represented as Δscore for NLI, NDI, and EMS, and JOA recovery rate (%) = (postoperative JOA score preoperative JOA score) / (17 preoperative JOA score) × 100.^26^

### 3. Mass spectrometry analysis of cerebrospinal fluid samples

For protein identification and quantification of CSF samples obtained from patients who underwent intramedullary surgery, a standard TMT-MS protocol was employed.^27^ CSF samples collected pre-and postoperatively were cleared, aliquoted, and stored at −80□ for subsequent analysis. Proteins of CSF samples were denatured, carboxymethylated, and fully trypsinized. The digested peptide samples were then labeled using the TMT labeling reagents (TMTpro™ 16plex, Thermo Scientific) according to the manufacturer’s instructions. Subsequently, we desalted, concentrated, and separated TMT-labeled peptides using high-performance liquid chromatography (HPLC, Proxeon EASY-nano LC 1200 liquid chromatography pump, Thermo Fisher Scientific, pre-column 100 μm × 2 cm, Acclaim PepMapTM 100, analytical column ES803A, 2 μm, 100 Å, 75 μm × 50 cm, Thermo Fisher Scientific). Furthermore, the TMT-labeled peptides were analyzed using an Orbitrap Fusion mass spectrometer (Thermo Fisher Scientific, San Jose, CA, USA).

We performed a data-dependent acquisition with tandem mass spectrometry (MS/MS) spectra using a higher-energy collisional dissociation approach.^28^ To identify peptide sequences in the samples against the human UniProtKB/Swiss-Prot database (14/03/2022 released version), the raw data were processed using Software Proteome Discoverer (version 2.5, Thermo Scientific). Based on the intensity of the TMT tags, the relative abundance of each protein across different samples was computed. The quality of the TMT-MS analysis was controlled by protein coverage, peptide counts, and spectral counts.

### 4. Statistical analysis and mathematical modeling

#### Statistical analysis

In this research, we employed GraphPad Prism, version 9.4.1 (GraphPad Software) to perform various statistical analyses based on the types and nature of the variables. Unpaired t-tests, one-way ANOVA, and Pearson correlation analysis were employed to assess the data.

#### Integration of CSF proteomics data with clinical evaluation by LASSO penalized regression

The MS proteomics data were analyzed and interpreted with the clinical evaluation using LASSO penalized regression. The LASSO regression analysis was conducted in R version 4.1.2 (R Foundation for Statistical Computing). The "glmnet" package was employed for identifying factors correlated with recovery rate and modeling.^29^ The TMT-MS proteomics and clinical evaluation data were cleaned, normalized, transformed, and filtered out for outliers, missing values, or low-quality data. Subsequently, a feature selection was performed on the selected dataset using regression analysis by placing a LASSO penalty on the regression coefficients to shrink unimportant coefficients to zero. The Lasso regression model was further trained to fit with a selected set of features by setting hyperparameters, including the regularization parameter that balances the trade-off between model complexity and goodness of fit. Then, the performance of the trained models with different penalty and feature selections was assessed using metrics such as mean absolute error (MAE), root mean squared error (RMSE), and the coefficient of determination (R^2^). To obtain a robust estimate, ten-fold cross-validation was conducted,^30, 31^ enabling the selection of the best combination of protein variables. Finally, to evaluate the generalizability of the models, cross-validation was performed.

## Results

### Participants

This research collected data between August 2020 and March 2023 from 43 adult patients with intramedullary lesions who were admitted to the Neurospine group at the Department of Neurosurgery, Huashan Hospital (**Figure 1A**). A total of 86 CSF samples were collected from the patients both pre-and postoperatively. Detailed clinical information of all 43 patients is provided in **eTable 1**. The statistics for the major clinical characteristics of these patients are illustrated in **Figure 1B**. Of these cases, 23 were women (54%) and 20 were men (47%), with a mean age of 42.8±11.8 years at the time of surgery. The location of lesions varied from C1 to T12 vertebral level based on MR images, with a distribution of 22 cervical (51%), 5 cervical-thoracic (12%), and 16 thoracic (37%) lesions, as illustrated in **Figure 1C**.

**Figure 1.**
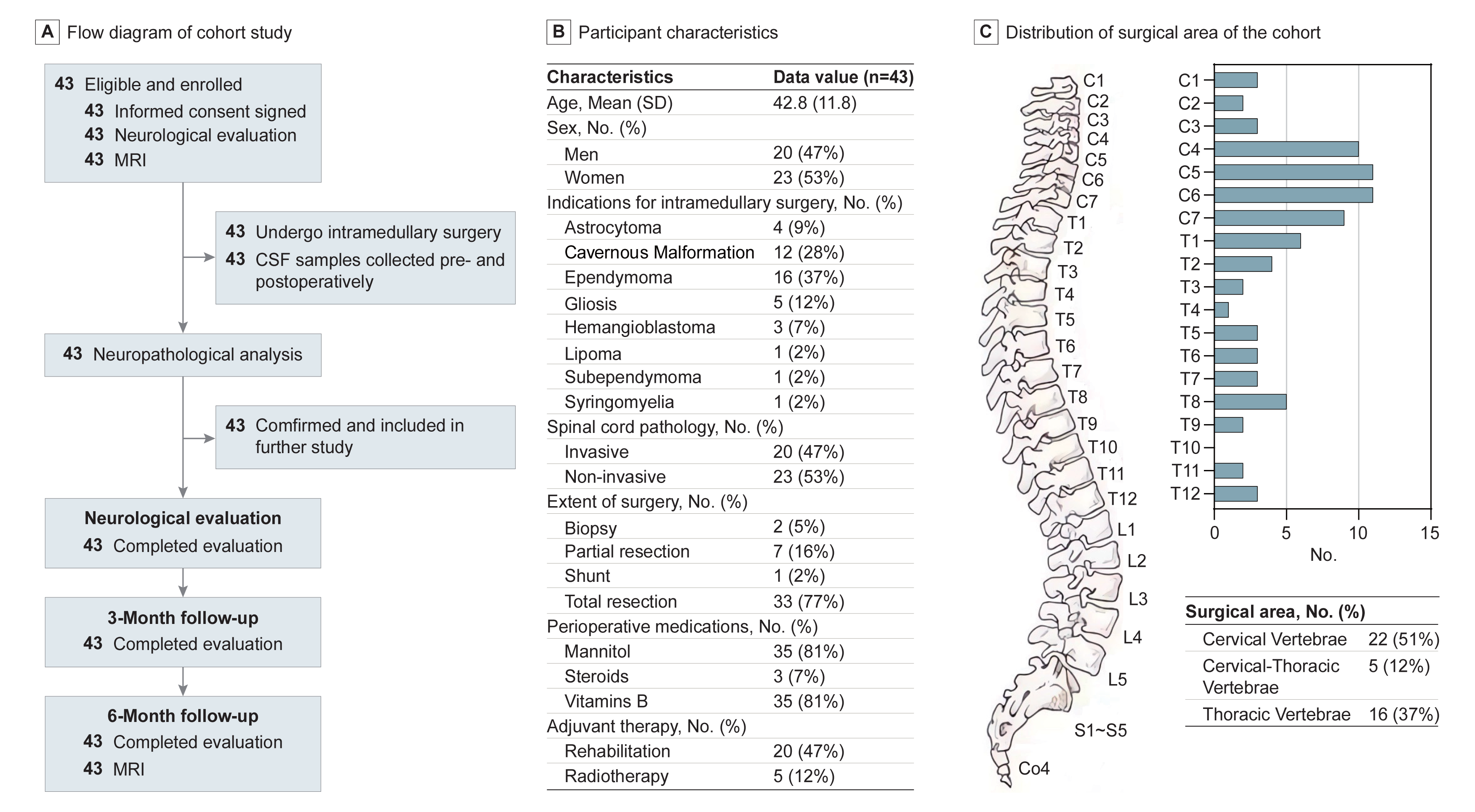
Flow Diagram of Cohort Research and Participant Characteristics. **A**. Flow diagram of the research. **B**. Participant characteristics **C**. The distribution of the surgical area of the cohort. The bars depict the accumulative affected spinal cord at every vertebral level. The distribution is presented in percentage according to the spinal vertebral with a cervical predominant.

### Surgical treatment and pathology

Total gross resections were achieved in 77% of the patients, while 16% underwent partial removal. Biopsy for histological diagnosis was performed in 5% of the patients. The pathological analysis identified 1 syringomyelia (2%), 1 lipoma (2%), 1 subependymoma (2%), 3 hemangioblastomas (7%), astrocytomas (9%), 5 gliosis (12%), 12 cavernous malformations (28%), and 16 ependymomas (37%) (**Figure 1B**). These distributions are consistent with previous reports in the literature.^3, 32^ Twenty cases (47%) were classified as invasive, with a histological diagnosis of astrocytoma or ependymoma according to the World Health Organization (WHO) classification of lesions in the central nervous system.^33, 34^ Postoperative treatment included using mannitol and vitamin B, which were widely administered, while only three patients received steroid therapies. Five patients underwent postoperative radiation therapy, and 20 patients received rehabilitation treatments, including hyperbaric oxygen therapy (**Figure 1B, eTable 1**).

### Follow-up study

All patients completed two postoperative follow-up visits at the 3rd and 6th months. We comprehensively evaluated the spinal function status of the patients using assessments such as the ASIA impairment scales, NDI, NLI, EMS, and JOA scores. At the 3rd and 6th month follow-ups, 23 patients demonstrated stable or improved spinal function, while 20 experienced deterioration. This trend was consistent across NDI, NLI, EMS, and JOA scores (**Figure 2**, **eTable 2**, **eFigure 1**). However, there was minimal difference in the AIS scales before and after surgery. **Figure 2A** illustrates the time course of the spinal function of all patients, with baseline injury levels ranging from C1 to L2 before the operation. The observed changes were consistent with the follow-up results at the 3rd and 6th months, indicating no significant differences. We employed JOA scoring to assess motor, sensory, and reflex functions in the upper and lower extremities (**Figure 2B**). The mean JOA score before surgery was 12.9, exhibiting considerable variations among patients (range: −8 to +3). No statistically significant difference was observed between the scores of the 3rd and 6th month follow-ups. These findings demonstrate that the functional status of patients remained stable during this period.

**Figure 2.**
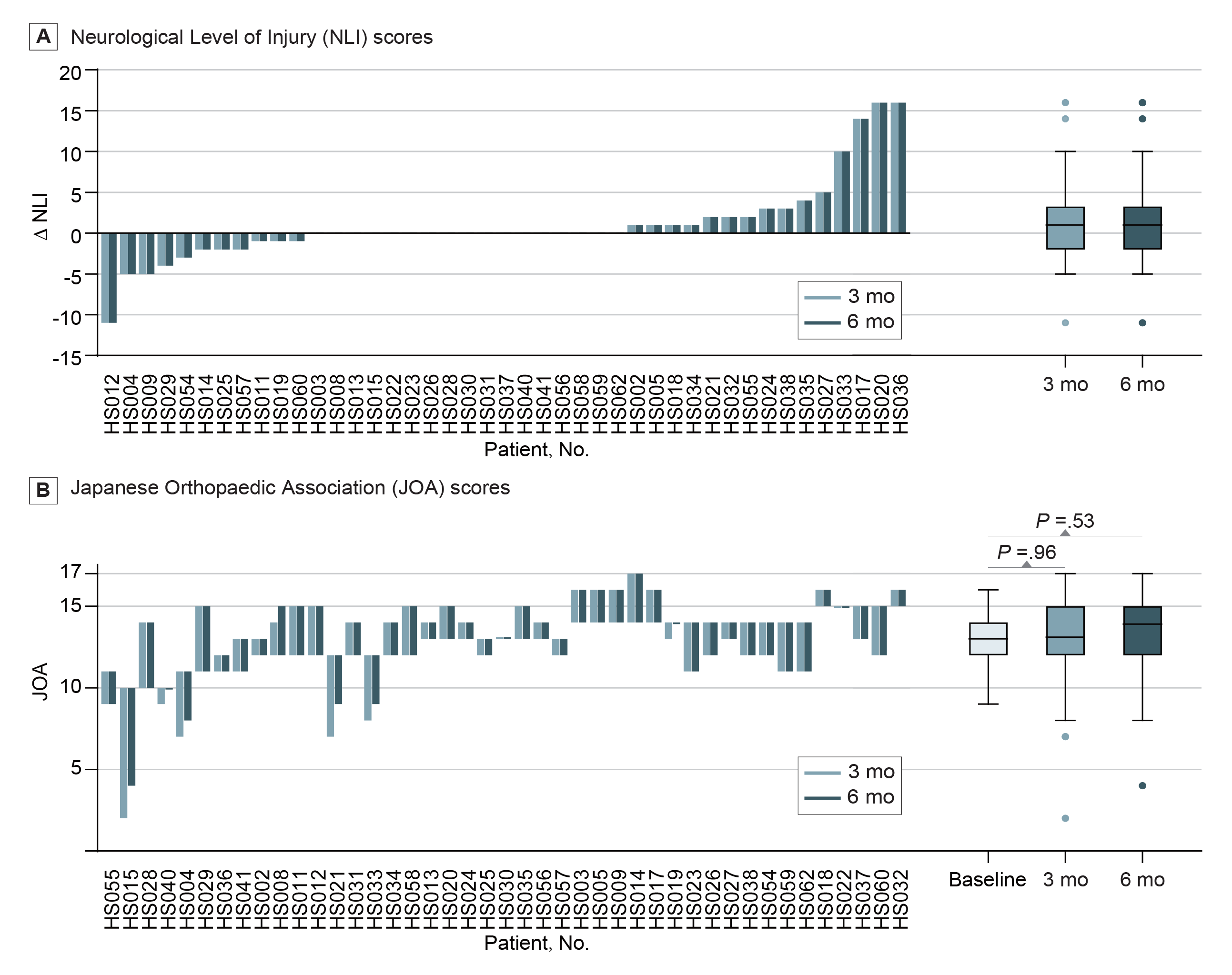
Neurological Assessment with Neurological Level of Injury (NLI) and Japanese Orthopaedic Association (JOA) Scores. **A**. NLI scores. The time course of spinal function was evaluated using NLI scores from baseline to the 3rd and 6th month postsurgery time points. The change in follow-up results at the 3rd and 6th months postoperatively was consistent, and there were no statistically significant differences between them. **B**. JOA scores. The time course of spinal function was evaluated using JOA scores. The change in JOA score varied from −8 to +3. There was no statistically significant difference between the 3rd and 6th month follow-up scores.

To examine the relationship between various clinical variables and functional changes at the 3rd month follow-up, we performed correlation analysis (**Table 1**). The analysis indicated no statistically significant correlation with factors such as age, pathology, lesion size, the extent of surgery, sex, perioperative medication, surgical area, and adjuvant therapy, as evaluated by the change in NDI and NLI scales. The steroid medication helped just a small percentage of individuals recover. Notably, lesion size demonstrated a strong correlation with the recovery rate of the JOA score, indicating that the JOA score recovery rate may be more sensitive in assessing functional improvement in these patients. In addition, a significant association was observed in pathological results (noninvasive versus invasive), with functional prognosis based on the changes in EMS scale and JOA score, with *P* < .05. We employed the JOA recovery rate and subgrouped patients with noninvasive versus invasive pathological characteristics for further analysis based on these findings.

**Table 1.** Correlation Analysis of Clinical Variables with 3-month Follow-up.

| Factors | P value |  |  |  |  |
| --- | --- | --- | --- | --- | --- |
|  | Δ NLI | Δ NDI | Δ EMS | Δ JOA | JOA recovery rate |
| Demographic |  |  |  |  |  |
| Age<sup>c</sup> | .25 | .97 | .70 | .41 | .74 |
| Sex<sup>a</sup> | .32 | .68 | .77 | .86 | .87 |
| Spinal cord pathology<sup>a</sup> | .32 | .69 | .02 | .03 | .006 |
| Size<sup>c</sup> | .60 | .27 | .12 | .007 | .005 |
| Extent of surgery<sup>b</sup> | .97 | .15 | .87 | .77 | .90 |
| Surgical area<sup>b</sup> (Number of segments<sup>b</sup>) | .18 (.95) | .95 (.88) | .82 (.50) | .63 (.35) | .88 (.15) |
| Perioperative medications |  |  |  |  |  |
| Mannitol<sup>a</sup> | .07 | .12 | .90 | .32 | .17 |
| Steroids<sup>a</sup> | .29 | .19 | .001 | <.001 | .01 |
| Vitamin B<sup>a</sup> | .32 | .31 | .73 | .64 | .43 |
| Adjuvant therapy |  |  |  |  |  |
| Rehabilitation<sup>a</sup> | .56 | .93 | .11 | .28 | .07 |
| Radiotherapy<sup>a</sup> | .63 | .06 | .10 | .07 | .21 |

<sup>a</sup> Categorical variables were compared using an unpaired t-test.
<sup>b</sup> Ordinal categorical variables were compared using one-way ANOVA.
<sup>c</sup> Continuous variables were compared using Pearson correlation analysis.

### CSF proteomics analysis, modeling, and biomarker identification

We successfully quantified more than 3,800 proteins by applying TMT-MS, with an average of 2,191 proteins quantified per CSF sample, and there was no considerable difference in the number of proteins detected (**Figure 3A**). The completeness of TMT was 100% for 1,152 proteins (30%), 75% for 1,558 proteins (40%), and 50% for 2,022 proteins (52%) (**Figure 3A**). Quantified protein intensities spanned over six orders of magnitude. Notably, the top ten most abundant proteins accounted for 25% of the total protein intensity in the entire dataset of 3,870 proteins (**Figure 3B**). Additionally, we discovered that 570 and 648 proteins from preoperative and postoperative samples, respectively, were significantly correlated with the JOA recovery rate (*P* < .05), with Pearson correlation coefficients ranging from −0.62 to 0.57 for the preoperative samples and −0.69 to 0.48 for the postoperative samples (**Figure 3C**). Then, we generated a heat map with the ratio of each protein from each patient (**Figure 3D**),^35^ and used a hierarchical clustering algorithm to group the proteins. The invasive and noninvasive samples spontaneously clustered into two groups, which were further sorted based on lesion sizes. However, no specific protein emerged as a priority for further validation or investigation of its potential association with spinal function recovery.

**Figure 3.**
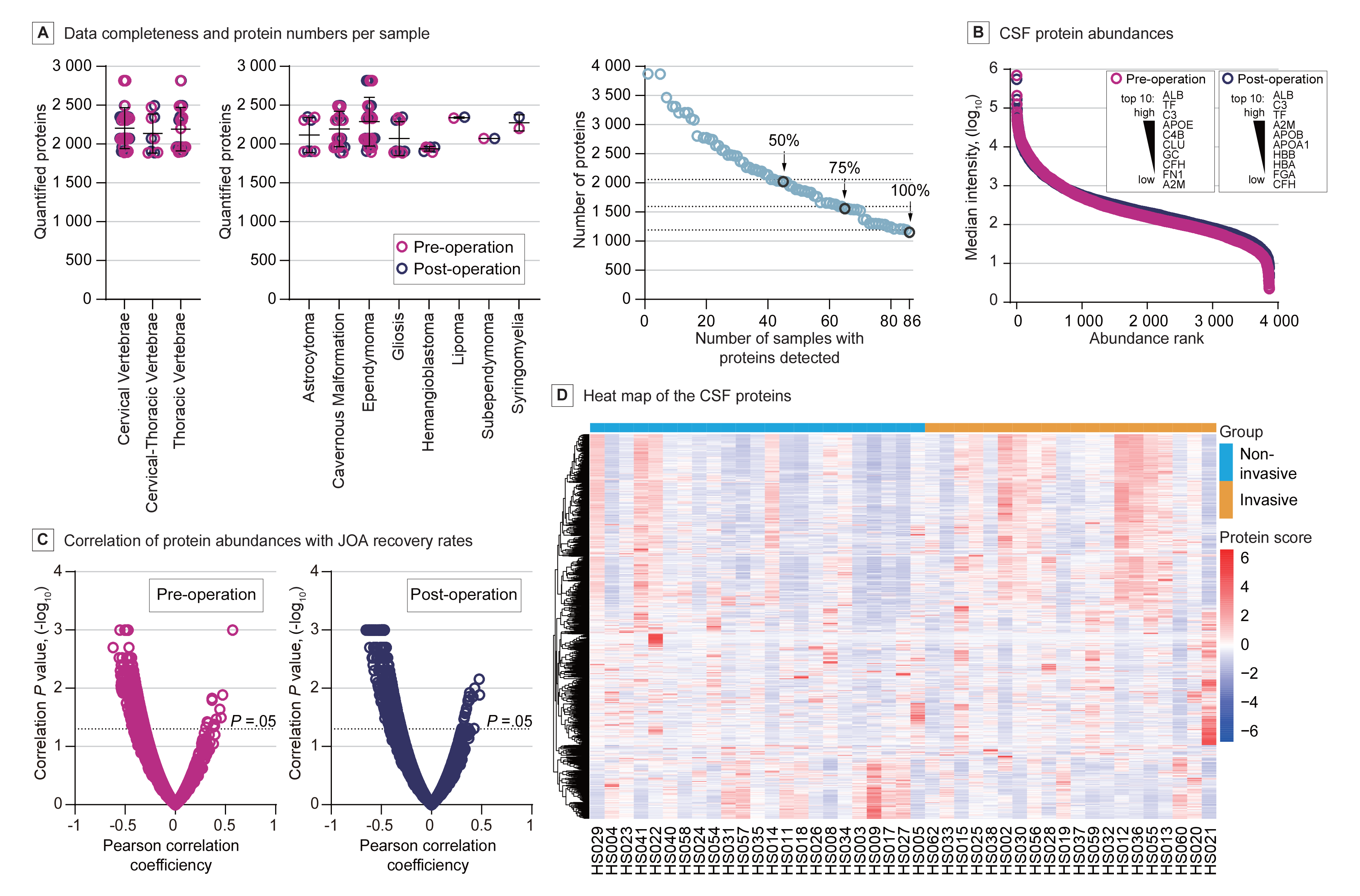
Proteomic Analysis of Cerebrospinal Fluid. **A**. Data completeness and protein numbers per sample. Horizontal lines show the mean and error bars for ± SD. The number of proteins in each sample was grouped based on their surgical area (left panel) or pathological analysis (middle panel) and shown accordingly. The number of proteins in the datasets (Y-axis) versus the minimum number of samples in which each protein had been quantified (X-axis) was plotted and shown as a data completeness curve (right panel). The arrows indicate 50%, 75%, and 100% data completeness. **B**. CSF protein abundance. The median CSF protein abundance distribution was calculated from MS intensities of quantified peptides of each protein. The top ten most abundant proteins are highlighted. **C**. The correlation of protein abundance with JOA recovery rates. The dashed line indicates the *P* value of 0.05. **D**. Heat map of the CSF proteins. Each row represents one protein, and each column represents one sample. Proteins were grouped using a hierarchical clustering algorithm. Noninvasive and invasive samples clustered spontaneously into two groups, which were further sorted by lesion size.

Then, we employed feature selection reduction algorithms for data analysis and interpretation. Specifically, we used LASSO analysis on the total datasets and the subsets of normalized and transformed TMT-MS data for noninvasive or invasive data. To optimize the regularization strength parameter, lambda, we employed cross-validation and evaluated its performance using RMSE and MAE metrics. Additionally, to determine the importance of each protein in predicting recovery after spinal cord surgery, we assessed the model performance using the R^2^ metric and obtained the coefficient scores (**eTable3 and 4**). Notably, we observed that the LASSO models demonstrated improved performance when applied to the noninvasive and invasive sub-population datasets compared to the entire population dataset. The LASSO models trained on the noninvasive or invasive sub-datasets exhibited lower RMSE, MAE, and higher R^2^ compared to those trained on the entire dataset when the number of protein variables (features) was kept constant (**Figure 4A-C**).

**Figure 4.**
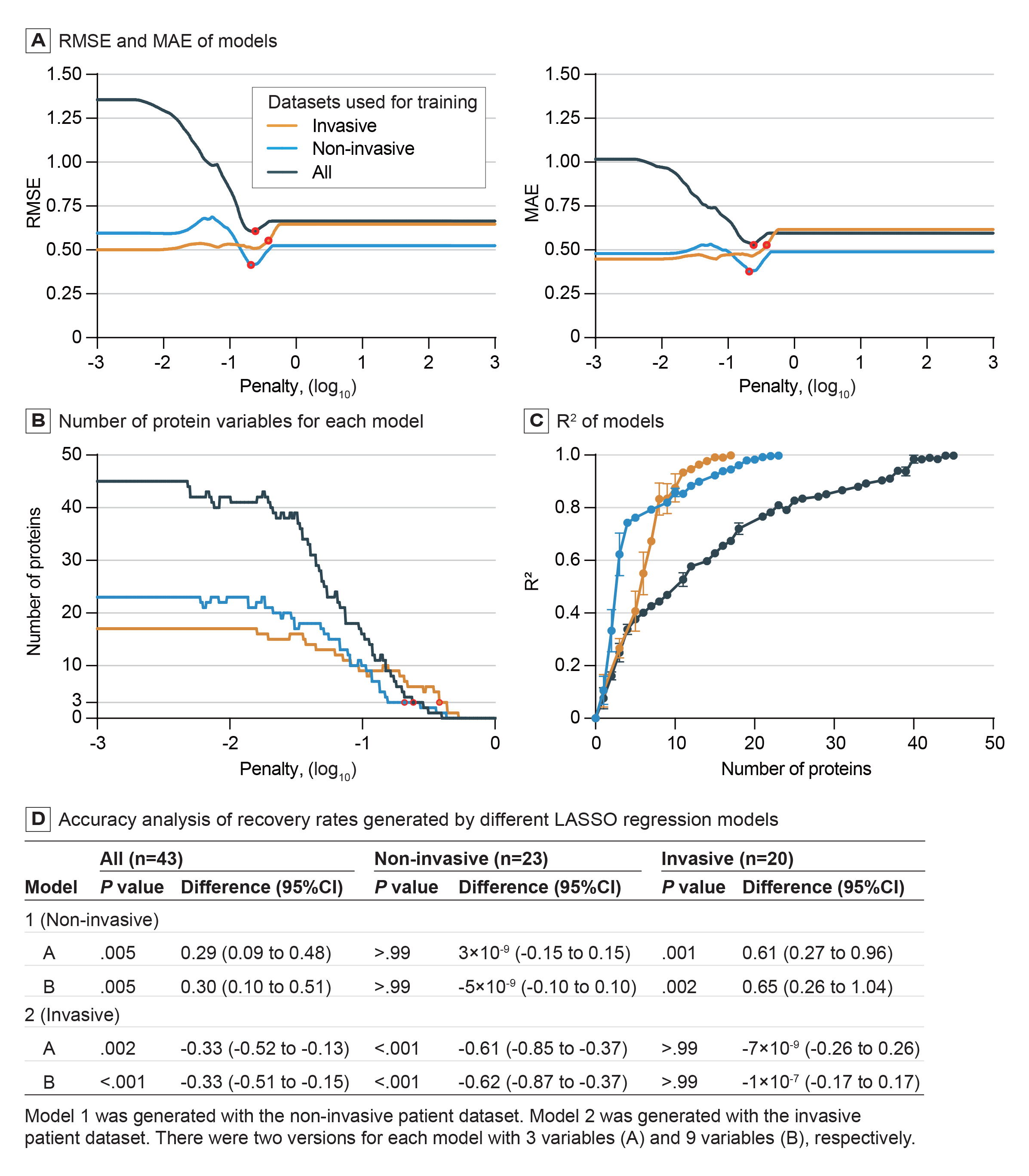
Parameters of LASSO Models Generated Using Different Training Datasets. **A**. RMSE (left panel) and MAE (right panel) of models. LASSO models were trained with 3 groups of datasets: the whole, the noninvasive, and the invasive datasets. Each model was trained with the same set of 1000 penalty values in the range of 10^−3^ to 10^3^ for each group. The points circled in red are the values of penalty when the model RMSE and MAE were the best ("All" curve penalty = 0.242, "noninvasive" curve penalty = 0.208, and ’invasive" curve penalty = 0.381). **B**. The number of protein variables for each model. Each group was trained with the same set of 1000 penalty parameter values in the range 10^−3^ to 10^3^ for each dataset. The maximum number of selected protein variables of the model trained with the whole dataset was 45, the maximum number of the noninvasive group was 24, and the maximum number of the invasive group was 17. For those models with a penalty parameter value greater than 1, the number of protein variables was 0 and was omitted from this figure. **C**. R^2^ of models. The number of protein variables selected (X-axis) versus the mean ± SD of R^2^ (Y-axis) was plotted. **D**. Accuracy analysis of recovery rates generated by different LASSO regression models. Model 1 was generated using the noninvasive patient dataset. Model 2 was generated using the invasive patient dataset. There were two versions for each model with 3 variables (A) and 9 variables (B), respectively. Analyses were conducted using paired t-tests with two-tailed *P* value and difference (95% CI).

We generated four models using noninvasive or invasive sub-datasets, consisting of 3 or 9 protein variables, respectively. These models were chosen based on their optimal MAE, RMSE, and R^2^ metrics among all feasible variable combinations. We analyzed the accuracy of recovery rates generated by these models. There was no significant difference between the recovery rates generated by the two models of the noninvasive or invasive populations compared to the original JOA recovery rates, as demonstrated in **Figure 4D**. However, we observed a significant difference between the generated recovery rates of the invasive populations by the two models trained on the noninvasive dataset and the original JOA recovery rates, and vice versa (*P* < .001) (Figure 4D). These findings indicate a mutual exclusivity between the invasive and the noninvasive sub-population.

To gain insights into the underlying biological mechanisms associated with the results of the LASSO models, we conducted pathway analysis and protein–protein interaction analysis (**eFigure 2B-C**). For the entire dataset, we identified 62 proteins with a variable importance score greater than 0. In the noninvasive group, 30 proteins exhibited a score greater than 0 (**eTable3**), of which 18 proteins scored 50 or higher. In this group, CTSG (95 points), HAGH (91 points), and PLIN4 (85 points) were the top three proteins. Similarly, 21 proteins exhibited a score greater than 0 in the invasive group (eTable4), with 14 proteins scoring 50 or higher. In this group, KLK11 (90 points), MFAP4 (86 points), and DNAJC3 (86 points) were the top three proteins. No protein was found shared with all three groups, as demonstrated in **eFigure 2A**. Six proteins overlapped between the entire dataset group and the noninvasive group (5.8%), while 3 proteins overlapped between the entire dataset group and the invasive group (2.9%). Notably, only one protein, MYBPC1 (Q00872), was shared by both the noninvasive and invasive groups, indicating distinct regulatory mechanisms underlying the noninvasive and invasive subgroups.

In the noninvasive group, enrichment analysis^36^ using the STRING protein–protein interaction database revealed a highly interconnected network of proteins (**eFigure 2B**). This indicates that these proteins work together (30 nodes/10 edges observed vs. 4 edges expected; mean node degree, 0.667; PPI enrichment significance, *P* = .003). However, in the invasive group, a loosely interconnected network of proteins was observed (14 nodes/1 edge observed vs. 0 edges expected; mean node degree, 0.143; PPI enrichment significance, *P* = .40). This indicates more diverse characteristics of this subgroup. In our future research, the validation of potential regulatory mechanisms involving these proteins will be conducted.

## Discussion

This cohort research encompasses clinically common intramedullary lesions, including vascular lesions (such as cavernous malformations and hemangioblastoma), neoplastic lesions (such as ependymoma and astrocytoma), as well as gliosis and syringomyelia. This single-centered observational research benefited from a stable surgical team, which helped minimize confounding variables that could affect clinical outcomes. Thus, this research offers an opportunity to observe a stronger correlation between clinical variables and surgical recovery. The findings of this research highlight a significant correlation between the underlying pathological characteristics and lesion sizes and neurological function recovery after intramedullary spinal cord surgery. These results are consistent with studies conducted by other medical teams, which reveal that the prognosis of patients largely depends on the invasiveness of the tumors ^3, 37–39^. Interestingly, we observed that patients with noninvasive tumors can be grouped with those with nontumorous lesions, demonstrating a shared characteristic among these cases. This observation motivates further investigations into the underlying biological regulatory mechanisms that contribute to the recovery of spinal cord impairment across these clinically diverse pathologies.

Valuable insights for postoperative medical intervention after intramedullary surgery have been provided by several retrospective cohort studies.^6, 39–41^ However, correlated biomedical research for new therapeutic methods still heavily depends on animal models, necessitating clearer and more direct connections to the clinical setting. CSF proteomics research serves as a potential bridge between these studies. Thus, it is crucial to conduct well-designed prospective cohort studies. In our cohort research, we scheduled all surgeries, enabling convenient access to pre-and postoperative CSF sample collection. We identified specific proteins or biomarkers associated with surgical intervention and the subsequent recovery process by comparing the protein profiles between these samples. This discovery is expected to enhance our understanding of the underlying biological mechanisms involved in spinal cord recovery and has the potential to identify therapeutic targets.

Consistent with previous investigations and reports from other research teams, this research demonstrates that despite advancements in intramedullary surgery, about 30% of patients attain a "favorable outcome" characterized by substantial improvement in neurological function. To detect significant differences in surgical outcomes associated with CSF features, conventional statistical estimation demonstrates the necessity of a cohort study involving a minimum of 100 patients, with 30 experiencing a "favorable outcome" and 70 not, ensuring 95% confidence and 80% power to detect a difference of at least 20%. However, due to the relatively low incidence of intramedullary lesions, recruiting sufficient patients for our single-centered prospective cohort research presents a significant challenge. Furthermore, evaluating recovery outcomes requires an extended follow-up period, which increases the risk of loss to follow-up and attrition. In addition, our results indicate that noninvasive and invasive patients may experience different factors influencing their recovery process, emphasizing the significance of considering the diverse pathological features when determining their recovery trajectory. Consequently, it becomes crucial to carefully subgroup the datasets to account for these variations.

To overcome the challenge of limited participants for each dataset group and the extensive proteomics data per sample, we employed LASSO regression modeling to examine and interpret CSF proteomics data alongside clinical evaluation across different patient groups. This research demonstrates that through feature selection, we can identify the most relevant protein sets for different patients with diverse outcomes, even within a small cohort. Additionally, LASSO regression modeling allows us to reduce the number of features (proteins) to a manageable size and prioritize the biomarkers to be investigated. In our research, LASSO regression models consisting of 3 or 9 CSF protein variables, along with 18 or 14 protein factors exhibiting relatively high scores (≥ 50, see eTable 3–4), indicate a robust correlation with the recovery of noninvasive and invasive patients, respectively.

In-depth investigations on traumatic spinal cord injuries present challenges due to the diverse range of severity, location, and other factors. This makes it particularly difficult to establish prospective cohort research. However, considering the similarities between the recovery processes after intramedullary noninvasive lesion removal and spinal cord injury, our current CSF analysis combined with the postsurgical recovery cohort research holds significant implications. It offers valuable insights into underlying regulatory mechanisms that can contribute to the development of new therapies for patients with spinal cord injuries, in addition to those undergoing intramedullary lesion removal.

### Limitations

This research has several limitations that should be acknowledged. First, ethical regulations regarding the protection of patients’ interests prohibited us from scheduling CSF sampling at the 3rd and 6th month follow-up periods for self-comparison. This limitation restricts us from gaining further valuable insights into the recovery process. Second, recruiting more patients is essential to further validate and stabilize these models. For this purpose, our cohort research is still ongoing. Third, due to the significant correlation between the prognosis of patients with invasive lesions and tumor malignant characteristics, it is crucial to design more specific and focused cohort research to identify the factors contributing to the postsurgical recovery of this subgroup of patients.

## Conclusions

This cohort research identified pathological characteristics and lesion size as the primary factors influencing patient neurological function recovery. In addition, this research developed LASSO regression models with the identified CSF protein variables, offering the potential for employing these models in predicting the prognosis of intramedullary surgery for patients with noninvasive and invasive lesions, respectively.

## Data Availability

All data produced in the present study are available upon reasonable request to the authors.

## Funding

Dr. Jian Zhao is supported by grants 2022YFA1105003 and 2018YFA0107903 from National Key Research and Development Program of China; grant YDZX20213100001684 from Shanghai Science and Technology Development Foundation; grant 31871408 from National Natural Science Foundation of China. Dr. Shixin Gu is supported by grant 81870970 from National Natural Science Foundation of China; grant 21511104506 from Science and Technology Commission of Shanghai Municipality. Dr. Shufei Ge is supported by grant 21010502500 from Science and Technology Commission of Shanghai Municipality; the startup fund of ShanghaiTech University.

### Author contributions

Drs Jian Zhao and Shixin Gu had full access to all the data in the study and take responsibility for the integrity of the data and the accuracy of the data analysis. Weihao Di, Wentao Gu, and Xike Peng served as co-first authors and contributed equally to this work.

Concepts, design and supervision: Jian Zhao, Shixin Gu. Acquisition, analysis, interpretation of data and drafting of the manuscript: All authors. Statistical analysis: Shufei Ge, Weihao Di, Xike Peng. Administrative, technical, or material support: Wentao Gu, Jiajun Shou, Jingyue Wang, Ying Wang, Na Guo, Wei Zhu.

## REFERENCES

1. Ostrom QT, Cioffi G, Waite K, Kruchko C, Barnholtz-Sloan JS. CBTRUS Statistical Report: Primary Brain and Other Central Nervous System Tumors Diagnosed in the United States in 2014-2018. Neuro Oncol. Oct 5 2021;23(12 Suppl 2):iii1–iii105. doi:10.1093/neuonc/noab200

2. Schellinger KA, Propp JM, Villano JL, McCarthy BJ. Descriptive epidemiology of primary spinal cord tumors. J Neurooncol. Apr 2008;87(2):173–9. doi:10.1007/s11060-007-9507-z

3. Endo T, Inoue T, Mizuno M, et al. Current Trends in the Surgical Management of Intramedullary Tumors: A Multicenter Study of 1,033 Patients by the Neurospinal Society of Japan. Neurospine. Jun 2022;19(2):441–452. doi:10.14245/ns.2244156.078

4. Harrop JS, Ganju A, Groff M, Bilsky M. Primary intramedullary tumors of the spinal cord. Spine (Phila Pa 1976). Oct 15 2009;34(22 Suppl):S69–77. doi:10.1097/BRS.0b013e3181b95c6f

5. Juthani RG, Bilsky MH, Vogelbaum MA. Current Management and Treatment Modalities for Intramedullary Spinal Cord Tumors. Curr Treat Options Oncol. Aug 2015;16(8):39. doi:10.1007/s11864-015-0358-0

6. Ottenhausen M, Ntoulias G, Bodhinayake I, et al. Intradural spinal tumors in adults-update on management and outcome. Neurosurg Rev. Jun 2019;42(2):371–388. doi:10.1007/s10143-018-0957-x

7. Roche S, Gabelle A, Lehmann S. Clinical proteomics of the cerebrospinal fluid: Towards the discovery of new biomarkers. Proteomics Clin Appl. Mar 2008;2(3):428–36. doi:10.1002/prca.200780040

8. Teunissen CE, Verheul C, Willemse EAJ. The use of cerebrospinal fluid in biomarker studies. Handb Clin Neurol. 2017;146:3–20. doi:10.1016/b978-0-12-804279-3.00001-0

9. Hühmer AF, Biringer RG, Amato H, Fonteh AN, Harrington MG. Protein analysis in human cerebrospinal fluid: Physiological aspects, current progress and future challenges. Dis Markers. 2006;22(1-2):3–26. doi:10.1155/2006/158797

10. Li J, Van Vranken JG, Pontano Vaites L, et al. TMTpro reagents: a set of isobaric labeling mass tags enables simultaneous proteome-wide measurements across 16 samples. Nat Methods. Apr 2020;17(4):399–404. doi:10.1038/s41592-020-0781-4

11. Liu D, Yang S, Kavdia K, et al. Deep Profiling of Microgram-Scale Proteome by Tandem Mass Tag Mass Spectrometry. J Proteome Res. Jan 1 2021;20(1):337–345. doi:10.1021/acs.jproteome.0c00426

12. Neagu AN, Jayathirtha M, Baxter E, Donnelly M, Petre BA, Darie CC. Applications of Tandem Mass Spectrometry (MS/MS) in Protein Analysis for Biomedical Research. Molecules. Apr 8 2022;27(8)doi:10.3390/molecules27082411

13. Valdés A, Bergström Lind S. Mass Spectrometry-Based Analysis of Time-Resolved Proteome Quantification. Proteomics. May 2020;20(9):e1800425. doi:10.1002/pmic.201800425

14. Dunkler D, Sánchez-Cabo F, Heinze G. Statistical analysis principles for Omics data. Methods Mol Biol. 2011;719:113–31. doi:10.1007/978-1-61779-027-0_5

15. Fan J, Han F, Liu H. Challenges of big data analysis. National science review. 2014;1(2):293–314.

16. Tibshirani R. Regression shrinkage and selection via the lasso. Journal of the Royal Statistical Society: Series B (Methodological). 1996;58(1):267–288.

17. Tibshirani R. The lasso method for variable selection in the Cox model. Stat Med. Feb 28 1997;16(4):385–95. doi:10.1002/(sici)1097-0258(19970228)16:4<385::aid-sim380>3.0.co;2-3

18. Oyeyemi GM. <On performance of shrinkage methods–a Monte Carlo Study.pdf>. 2015;doi:10.5923/j.statistics.20150502.04

19. Wang P, Chen S, Yang S. Recent advances on penalized regression models for biological data. Mathematics. 2022;10(19):3695.

20. Kashyap H, Ahmed HA, Hoque N, Roy S, Bhattacharyya DK. Big data analytics in bioinformatics: A machine learning perspective. arXiv preprint arXiv:150605101. 2015;

21. Roberts TT, Leonard GR, Cepela DJ. Classifications In Brief: American Spinal Injury Association (ASIA) Impairment Scale. Clin Orthop Relat Res. May 2017;475(5):1499–1504. doi:10.1007/s11999-016-5133-4

22. Vernon H, Mior S. The Neck Disability Index: a study of reliability and validity. J Manipulative Physiol Ther. Sep 1991;14(7):409–15.

23. Vitzthum HE, Dalitz K. Analysis of five specific scores for cervical spondylogenic myelopathy. Eur Spine J. Dec 2007;16(12):2096–103. doi:10.1007/s00586-007-0512-x

24. Benzel EC, Lancon J, Kesterson L, Hadden T. Cervical laminectomy and dentate ligament section for cervical spondylotic myelopathy. J Spinal Disord. Sep 1991;4(3):286–95. doi:10.1097/00002517-199109000-00005

25. Hirabayashi K, Miyakawa J, Satomi K, Maruyama T, Wakano K. Operative results and postoperative progression of ossification among patients with ossification of cervical posterior longitudinal ligament. Spine (Phila Pa 1976). Jul-Aug 1981;6(4):354–64. doi:10.1097/00007632-198107000-00005

26. Tetreault L, Kopjar B, Nouri A, et al. The modified Japanese Orthopaedic Association scale: establishing criteria for mild, moderate and severe impairment in patients with degenerative cervical myelopathy. Eur Spine J. Jan 2017;26(1):78–84. doi:10.1007/s00586-016-4660-8

27. Thompson A, Wölmer N, Koncarevic S, et al. TMTpro: Design, Synthesis, and Initial Evaluation of a Proline-Based Isobaric 16-Plex Tandem Mass Tag Reagent Set. Anal Chem. Dec 17 2019;91(24):15941–15950. doi:10.1021/acs.analchem.9b04474

28. Jora M, Burns AP, Ross RL, et al. Differentiating Positional Isomers of Nucleoside Modifications by Higher-Energy Collisional Dissociation Mass Spectrometry (HCD MS). J Am Soc Mass Spectrom. Aug 2018;29(8):1745–1756. doi:10.1007/s13361-018-1999-6

29. Friedman JH, Hastie T, Tibshirani R. Regularization Paths for Generalized Linear Models via Coordinate Descent. Journal of Statistical Software. 02/02 2010;33(1):1–22. doi:10.18637/jss.v033.i01

30. Kohavi R. A study of cross-validation and bootstrap for accuracy estimation and model selection. presented at: Proceedings of the 14th international joint conference on Artificial intelligence - Volume 2; 1995; Montreal, Quebec, Canada.

31. Berrar D. Cross-Validation. 2019.

32. Boström A, Kanther NC, Grote A, Boström J. Management and outcome in adult intramedullary spinal cord tumours: a 20-year single institution experience. BMC Res Notes. Dec 15 2014;7:908. doi:10.1186/1756-0500-7-908

33. Louis DN, Perry A, Wesseling P, et al. The 2021 WHO Classification of Tumors of the Central Nervous System: a summary. Neuro Oncol. Aug 2 2021;23(8):1231–1251. doi:10.1093/neuonc/noab106

34. Smith HL, Wadhwani N, Horbinski C. Major Features of the 2021 WHO Classification of CNS Tumors. Neurotherapeutics. May 16 2022;doi:10.1007/s13311-022-01249-0

35. Babicki S, Arndt D, Marcu A, et al. Heatmapper: web-enabled heat mapping for all. Nucleic Acids Res. Jul 8 2016;44(W1):W147–53. doi:10.1093/nar/gkw419

36. Szklarczyk D, Gable AL, Lyon D, et al. STRING v11: protein-protein association networks with increased coverage, supporting functional discovery in genome-wide experimental datasets. Nucleic Acids Res. Jan 8 2019;47(D1):D607–d613. doi:10.1093/nar/gky1131

37. Knafo S, Aghakhani N, David P, Parker F. Management of intramedullary spinal cord tumors: A single-center experience of 247 patients. Rev Neurol (Paris). May 2021;177(5):508–514. doi:10.1016/j.neurol.2020.07.014

38. Hersh AM, Patel J, Pennington Z, et al. Perioperative outcomes and survival after surgery for intramedullary spinal cord tumors: a single-institution series of 302 patients. J Neurosurg Spine. Feb 25 2022:1–11. doi:10.3171/2022.1.spine211235

39. Hussain I, Parker WE, Barzilai O, Bilsky MH. Surgical Management of Intramedullary Spinal Cord Tumors. Neurosurg Clin N Am. Apr 2020;31(2):237–249. doi:10.1016/j.nec.2019.12.004

40. Butenschoen VM, Nehiba A, Meyer B, Wostrack M. Neuropathic pain after spinal intradural benign tumor surgery: an underestimated complication? Neurosurg Rev. Aug 2022;45(4):2681–2687. doi:10.1007/s10143-022-01775-7

41. Costăchescu B, Niculescu AG, Iliescu BF, Dabija MG, Grumezescu AM, Rotariu D. Current and Emerging Approaches for Spine Tumor Treatment. Int J Mol Sci. Dec 10 2022;23(24)doi:10.3390/ijms232415680

