## Supplemental tables and figures for "Identification of Cerebrospinal Fluid Proteomic Signatures Associated with Intramedullary Surgery Outcome"

**Supplements**

**Contents**

Supplement 1 - eTable 1. Detailed Characteristics of Patients.

Supplement 2 - eTable 2. Detailed Neurological Assessment of Patients.

Supplement 3 - eFigure 1. Neurological Assessment with EMS and NDI.

Supplement 4 - eTable 3. Protein Selected by LASSO Regression Analysis with Noninvasive Subdatasets

Supplement 5 - eTable 4. Protein Selected by LASSO Regression Analysis with Invasive Subdatasets

Supplement 6 - eFigure 2. Biological Indications of Model Protein Variables

Supplement 7 - eMethods

**eTables**

#### eTable 1. Detailed Characteristics of Patients

| Patient, NO. | Age (provided as a range, per MedRxiv policy) | Sex | Indications for intramedullary surgery (Size*) | Spinal cord pathology | Extent of surgery | Surgical area (Number of segments) | Perioperative medications | Adjuvant therapy |
|---|---|---|---|---|---|---|---|---|
| | | | | | | | Mnt / S / V | Rhb / Rt |
| HS002 | 36-40 | M | Astrocytoma (15.9) | Invasive | Biopsy | C4 (1) | - / - / - | - / + |
| HS003 | 36-40 | W | CM (5.2) | Non-Invasive | Total resection | T6 (1) | - / - / - | - / - |
| HS004 | 31-35 | W | CM (NA) | Non-Invasive | Total resection | T8-9 (2) | - / - / - | + / - |
| HS005 | 36-40 | W | Gliosis (4.2) | Non-Invasive | Partial resection | C5 (1) | - / - / + | - / - |
| HS008 | 31-35 | M | Gliosis (6.0) | Non-Invasive | Biopsy | C1 (1) | - / - / + | - / - |
| HS009 | 31-35 | M | CM (4.4) | Non-Invasive | Total resection | C7-T1 (2) | + / - / + | - / - |
| HS011 | 36-40 | M | Gliosis (6.9) | Non-Invasive | Partial resection | C3-4 (2) | + / - / + | - / - |
| HS012 | 66-70 | W | EPN (10.4) | Invasive | Total resection | T1-2 (2) | - / - / + | - / - |
| HS013 | 36-40 | M | Astrocytoma (9.4) | Invasive | Partial resection | T11-12 (2) | + / - / + | - / + |
| HS014 | 31-35 | M | HB (7.2) | Non-Invasive | Total resection | C1 (1) | + / - / + | - / - |
| HS015 | 31-35 | W | Astrocytoma (17.7) | Invasive | Partial resection | C4-5 (2) | + / + / + | - / + |
| HS017 | 46-50 | W | CM (4.4) | Non-Invasive | Total resection | T7-8 (2) | + / - / + | - / - |
| HS018 | 46-50 | W | Gliosis (6.8) | Non-Invasive | Total resection | C7-T1 (2) | + / - / + | - / - |
| HS019 | 31-35 | M | Astrocytoma (13.3) | Invasive | Partial resection | C5-7 (3) | - / - / + | - / + |
| HS020 | 51-55 | W | EPN (6.3) | Invasive | Total resection | C4 (1) | + / - / + | - / - |
| HS021 | 41-45 | W | EPN (5.6) | Invasive | Total resection | C4-5 (2) | + / + / + | + / - |
| HS022 | 31-35 | W | Syringomyelia (12.7) | Non-Invasive | Shunt | T12 (1) | + / - / + | + / - |
| HS023 | 41-45 | W | Lipoma (25.9) | Non-Invasive | Partial resection | C7-T2 (3) | + / - / + | + / - |
| HS024 | 56-60 | W | Gliosis (10.0) | Non-Invasive | Partial resection | C2-4 (3) | + / - / + | - / - |
| HS025 | 56-60 | M | EPN (17.7) | Invasive | Total resection | C4-6 (3) | + / - / + | - / - |
| HS026 | 36-40 | M | CM (6.3) | Non-Invasive | Total resection | T5-6 (2) | + / - / + | + / - |
| HS027 | 46-50 | W | CM (4.3) | Non-Invasive | Total resection | T2-3 (2) | + / - / + | - / - |
| HS028 | 41-45 | M | EPN (14.2) | Invasive | Total resection | C6-T1 (3) | + / - / + | - / - |
| HS029 | 56-60 | W | CM (NA) | Non-Invasive | Total resection | T7-8 (2) | + / - / + | + / - |
| HS030 | 56-60 | M | EPN (14.2) | Invasive | Total resection | C5-6 (2) | + / - / + | + / - |
| HS031 | 26-30 | W | CM (8.1) | Non-Invasive | Total resection | C6 (1) | - / - / + | - / - |
| HS032 | 36-40 | M | EPN (10.4) | Invasive | Total resection | C5-6 (2) | + / - / - | + / - |
| HS033 | 41-45 | M | EPN (18.4) | Invasive | Total resection | C6-7 (2) | + / + / + | + / - |
| HS034 | 36-40 | M | CM (5.4) | Non-Invasive | Total resection | C1 (1) | + / - / + | + / - |
| HS035 | 51-55 | W | SEPN (7.4) | Non-Invasive | Total resection | C4-5 (2) | + / - / - | + / - |
| HS036 | 56-60 | W | EPN (10.1) | Invasive | Total resection | C6-7 (2) | + / - / - | + / - |
| HS037 | 16-20 | W | EPN (12.0) | Invasive | Total resection | C7-T3 (4) | + / - / - | + / - |
| HS038 | 56-60 | M | EPN (17.2) | Invasive | Total resection | C4-7 (4) | + / - / - | - / - |
| HS040 | 26-30 | M | CM (11.7) | Non-Invasive | Total resection | C5-6 (2) | + / - / + | - / - |
| HS041 | 41-45 | W | HB (13.4) | Non-Invasive | Total resection | T11-12 (2) | + / - / + | + / - |
| HS054 | 16-20 | W | HB (9.1) | Non-Invasive | Total resection | T9 (1) | + / - / + | - / - |
| HS055 | 55-60 | W | EPN (10.0) | Invasive | Total resection | T7-8 (2) | + / - / + | + / - |

| Patient, NO. | Age (provided as a range, per MedRxiv policy) | Sex | Indications for intramedullary surgery (Size*) | Spinal cord pathology | Extent of surgery | Surgical area (Number of segments) | Perioperative medications | Adjuvant therapy |
|---|---|---|---|---|---|---|---|---|
| | | | | | | | Mnt / S / V | Rhb / Rt |
| HS056 | 45-50 | W | EPN (14.2) | Invasive | Total resection | T5-6 (2) | + / - / + | + / - |
| HS057 | 45-50 | M | CM (7.4) | Non-Invasive | Total resection | T8 (1) | + / - / + | - / - |
| HS058 | 36-40 | M | CM (10.1) | Non-Invasive | Total resection | T4 (1) | + / - / + | + / - |
| HS059 | 65-70 | M | EPN (11.7) | Invasive | Total resection | C2-3 (2) | + / - / + | + / + |
| HS060 | 56-60 | W | EPN (8.4) | Invasive | Total resection | T5 (1) | + / - / + | + / - |
| HS062 | 41-45 | M | EPN (NA) | Invasive | Total resection | C4-6 (3) | + / - / + | + / - |

M, men; W, Women; CM, Cavernous Malformation; EPN, Ependymoma; HB, Hemangioblastoma; SEPN, Subependymoma; +, yes; -, No; Mnt, Mannitol; S, Steroids; V, Vitamins B; Rhb, Rehabilitation; Rt, Radiotherapy; *, Maximum diameter of the lesion on MRI horizontal plane, mm; NA, not available.

#### eTable 2. Detailed Neurological Assessment of Patients

| Patient, NO. | JOA | AIS | NLI | EMS | NDI |
|---|---|---|---|---|---|
| | B / 3 mo / 6 mo | B / 3 mo / 6 mo | B / 3 mo / 6 mo | B / 3 mo / 6 mo | B / 3 mo / 6 mo |
| HS002 | 12 / 13 / 13 | D / D / D | C3 / C4 / C4 | 14 / 14 / 14 | 2% / 12% / 12% |
| HS003 | 14 / 16 / 16 | D / D / D | T12 / T12 / T12 | 15 / 16 / 16 | 6% / 2% / 2% |
| HS004 | 11 / 7 / 8 | D / D / D | T12 / T7 / T7 | 14 / 9 / 10 | 2% / 16% / 14% |
| HS005 | 14 / 16 / 16 | D / D / D | C4 / C5 / C5 | 15 / 16 / 16 | 0 / 32% / 32% |
| HS008 | 12 / 14 / 15 | D / D / D | C1 / C1 / C1 | 14 / 16 / 16 | 2% / 2% / 2% |
| HS009 | 14 / 16 / 16 | D / D / D | T1 / C3 / C3 | 16 / 17 / 17 | 2% / 2% / 2% |
| HS011 | 12 / 15 / 15 | D / D / D | C4 / C3 / C3 | 13 / 16 / 16 | 2% / 2% / 2% |
| HS012 | 12 / 15 / 15 | D / D / D | L1 / T2 / T2 | 14 / 16 / 16 | 2% / 2% / 2% |
| HS013 | 13 / 14 / 14 | D / D / D | T12 / T12 / T12 | 15 / 16 / 16 | 2% / 2% / 2% |
| HS014 | 14 / 17 / 17 | D / D / D | C4 / C2 / C2 | 14 / 18 / 18 | 2% / 0 / 0 |
| HS015 | 10 / 2 / 4 | D / C / C | C4 / C4 / C4 | 9 / 7 / 8 | 12% / 20% / 20% |
| HS017 | 14 / 16 / 16 | D / D / D | C4 / T11 / T11 | 14 / 17 / 17 | 10% / 6% / 6% |
| HS018 | 15 / 16 / 16 | D / D / D | C1 / C2 / C2 | 16 / 16 / 16 | 18% / 20% / 20% |
| HS019 | 14 / 13 / 14 | D / D / D | C5 / C4 / C4 | 16 / 15 / 16 | 12% / 16% / 16% |
| HS020 | 13 / 15 / 15 | D / D / D | C3 / T12 / T12 | 15 / 14 / 14 | 30% / 8% / 8% |
| HS021 | 12 / 7 / 9 | D / D / D | C1 / C3 / C3 | 14 / 8 / 10 | 8% / 16% / 16% |
| HS022 | 15 / 15 / 15 | D / D / D | T11 / T11 / T11 | 15 / 16 / 16 | 8% / 2% / 2% |
| HS023 | 14 / 11 / 11 | D / D / D | T12 / T12 / T12 | 15 / 14 / 14 | 2% / 8% / 8% |
| HS024 | 13 / 14 / 14 | D / D / D | T12 / L3 / L3 | 14 / 14 / 14 | 2% / 2% / 2% |
| HS025 | 13 / 12 / 12 | D / D / D | C4 / C2 / C2 | 14 / 14 / 14 | 2% / 2% / 2% |
| HS026 | 14 / 12 / 12 | D / D / D | T6 / T6 / T6 | 15 / 15 / 15 | 2% / 4% / 4% |
| HS027 | 14 / 13 / 13 | D / D / D | C4 / T2 / T2 | 15 / 15 / 15 | 10% / 4% / 4% |
| HS028 | 10 / 14 / 14 | D / D / D | C4 / C4 / C4 | 13 / 16 / 16 | 10% / 2% / 2% |
| HS029 | 11 / 15 / 15 | D / D / D | L2 / T10 / T10 | 13 / 17 / 17 | 8% / 2% / 2% |
| HS030 | 13 / 13 / 13 | D / D / D | C4 / C4 / C4 | 14 / 16 / 16 | 8% / 2% / 2% |
| HS031 | 12 / 14 / 14 | D / D / D | C6 / C6 / C6 | 13 / 16 / 16 | 8% / 6% / 6% |
| HS032 | 16 / 15 / 15 | D / D / D | C2 / C4 / C4 | 16 / 15 / 15 | 30% / 8% / 8% |
| HS033 | 12 / 8 / 9 | D / D / D | C4 / T7 / T7 | 15 / 12 / 12 | 8% / 12% / 12% |
| HS034 | 12 / 14 / 14 | D / D / D | C1 / C2 / C2 | 15 / 15 / 15 | 22% / 12% / 12% |
| HS035 | 13 / 15 / 15 | D / D / D | C4 / T1 / T1 | 14 / 16 / 16 | 26% / 6% / 6% |
| HS036 | 11 / 12 / 12 | D / D / D | C2 / T11 / T11 | 10 / 16 / 16 | 22% / 10% / 10% |
| HS037 | 15 / 13 / 13 | D / D / D | T2 / T2 / T2 | 17 / 15 / 15 | 12% / 4% / 4% |
| HS038 | 14 / 12 / 12 | D / D / D | C1 / C4 / C4 | 16 / 15 / 15 | 24% / 32% / 32% |
| HS040 | 10 / 9 / 10 | D / D / D | C4 / C4 / C4 | 14 / 14 / 14 | 32% / 22% / 22% |
| HS041 | 11 / 13 / 13 | D / D / D | L1 / L1 / L1 | 12 / 13 / 13 | 12% / 12% / 12% |
| HS054 | 14 / 12 / 12 | D / D / D | L1 / T10 / T10 | 16 / 16 / 16 | 4% / 12% / 12% |
| HS055 | 9 / 11 / 11 | D / D / D | T9 / T11 / T11 | 10 / 11 / 11 | 14% / 12% / 12% |
| HS056 | 13 / 14 / 14 | D / D / D | T4 / T4 / T4 | 13 / 13 / 13 | 6% / 6% / 6% |
| HS057 | 13 / 12 / 12 | D / D / D | L1 / T11 / T11 | 14 / 15 / 15 | 16% / 14% / 14% |
| HS058 | 12 / 15 / 15 | D / D / D | T3 / T3 / T3 | 13 / 16 / 16 | 24% / 18% / 18% |
| HS059 | 14 / 11 / 11 | D / D / D | C2 / C2 / C2 | 15 / 11 / 11 | 12% / 26% / 26% |
| HS060 | 15 / 12 / 12 | D / D / D | T4 / T3 / T3 | 16 / 13 / 13 | 16% / 24% / 24% |
| HS062 | 14 / 11 / 11 | D / D / D | C3 / C3 / C3 | 14 / 12 / 12 | 10% / 22% / 22% |

B, Baseline; 3 mo, 3-month follow-up post-operation; 6 mo, 6-month follow-up post-operation; JOA, Japanese Orthopaedic Association; AIS, ASIA Impairment Scale; NLI, Neurological Level of Injury; EMS, European myelopathy score; NDI, Neck Disability Index.

#### eTable 3. Protein Selected by LASSO Regression Analysis with Noninvasive Subdatasets

| Uniprot ID | Protein name | Gene name | Score | Median (IQR) Pre-operation | Median (IQR) Post-operation | *P* value |
|---|---|---|---|---|---|---|
| P08311 | Cathepsin G | CTSG | 95 | 137.5 (200.2) | 595.1 (529.2) | <.001 |
| Q16775 | Hydroxyacylglutathione hydrolase, mitochondrial | HAGH | 91 | 247 (167.5) | 862 (997.5) | <.001 |
| Q96Q06 | Perilipin-4 | PLIN4 | 85 | 415 (238.1) | 1968.1 (2618.5) | <.001 |
| P35609 | Alpha-actinin-2 | ACTN2 | 73 | 1536.7 (1097.5) | 6206.6 (11684.7) | <.001 |
| P53634 | Dipeptidyl peptidase 1 | CTSC | 72 | 1122.5 (1558.1) | 408.1 (648.6) | <.001 |
| P16157 | Ankyrin-1 | ANK1 | 70 | 585.9 (866.6) | 1868.7 (3198.3) | <.001 |
| Q9UNZ2 | NSFL1 cofactor p47 | NSFL1C | 70 | 509.8 (668.5) | 1355 (1469.3) | <.001 |
| Q00872 | Myosin-binding protein C, slow-type | MYBPC1 | 67 | 534 (318.2) | 1949.8 (2942.8) | <.001 |
| P14780 | Matrix metalloproteinase-9 | MMP9 | 67 | 297.1 (203.5) | 1205.9 (1764.9) | <.001 |
| Q9Y6R7 | IgGFc-binding protein | FCGBP | 65 | 20604.2 (10551.1) | 8023.1 (5633.7) | <.001 |
| P02042 | Hemoglobin subunit delta | HBD | 63 | 1907.2 (4107.2) | 17289.3 (24858.2) | <.001 |
| P04003 | C4b-binding protein alpha chain | C4BPA | 62 | 3298 (2207) | 19143.2 (17097.2) | <.001 |
| P01833 | Polymeric immunoglobulin receptor | PIGR | 60 | 683.3 (424.5) | 1372.9 (1281.4) | <.001 |
| P14649 | Myosin light chain 6B | MYL6B | 59 | 652.9 (467) | 8433.9 (7691.6) | <.001 |
| Q14894 | Ketimine reductase mu-crystallin | CRYM | 59 | 434.7 (483.3) | 207.6 (258.2) | .005 |
| P46821 | Microtubule-associated protein 1B | MAP1B | 58 | 545 (779.9) | 972.6 (1928.8) | .03 |
| Q12913 | Receptor-type tyrosine-protein phosphatase eta | PTPRJ | 55 | 488 (806.9) | 421.3 (668.7) | .19 |
| Q8N436 | Inactive carboxypeptidase-like protein X2 | CPXM2 | 51 | 271.3 (452.6) | 158.4 (214.8) | <.001 |
| Q6YHK3 | CD109 antigen | CD109 | 49 | 1707.2 (1103.3) | 1212.9 (555.2) | <.001 |
| Q96SM3 | Probable carboxypeptidase X1 | CPXM1 | 48 | 115.95 (178.2) | 65.7 (58.625) | <.001 |
| Q6UWN8 | Serine protease inhibitor Kazal-type 6 | SPINK6 | 42 | 490.7 (769.1) | 120.7 (214.875) | <.001 |
| P00742 | Coagulation factor X | F10 | 38 | 3536.7 (2445.8) | 3167.4 (3100.8) | .14 |
| P26447 | Protein S100-A4 | S100A4 | 38 | 340.3 (480.3) | 3193.1 (3387.9) | <.001 |
| P81605 | Dermcidin | DCD | 37 | 788.4 (1448.9) | 760.4 (1029.8) | .07 |
| P35580 | Myosin-10 | MYH10 | 33 | 205.5 (280.7) | 349.4 (472.1) | <.001 |
| Q07507 | Dermatopontin | DPT | 30 | 582.1 (457.7) | 776.7 (618.925) | .002 |
| P50991 | T-complex protein 1 subunit delta | CCT4 | 27 | 208.4 (433.8) | 712.7 (1122.9) | <.001 |
| Q15084 | Protein disulfide-isomerase A6 | PDIA6 | 21 | 292.8 (420.6) | 269.8 (321.2) | .08 |
| Q13103 | Secreted phosphoprotein 24 | SPP2 | 21 | 322 (300.7) | 502.6 (605.7) | <.001 |
| P13611 | Versican core protein | VCAN | 10 | 4172 (4193.5) | 2391 (3051.5) | .04 |

#### eTable 4. Protein Selected by LASSO Regression Analysis with Invasive Subdatasets

| Uniprot ID | Protein name | Gene name | Score | Median (IQR) | | *P* value |
|---|---|---|---|---|---|---|
| | | | | Pre-operation | Post-operation | |
| Q9UBX7 | Kallikrein-11 | KLK11 | 90 | 500.6 (560.1) | 234.7 (285.4) | <.001 |
| P55083 | Microfibril-associated glycoprotein 4 | MFAP4 | 86 | 2625.7 (2046.3) | 726 (688.9) | <.001 |
| Q13217 | DnaJ homolog subfamily C member 3 | DNAJC3 | 86 | 573.5 (591.2) | 249.9 (221.1) | <.001 |
| P24593 | Insulin-like growth factor-binding protein 5 | IGFBP5 | 83 | 4437.5 (3539.8) | 1955.9 (1726.6) | <.001 |
| Q02809 | Procollagen-lysine,2-oxoglutarate 5-dioxygenase 1 | PLOD1 | 83 | 1174.5 (842.5) | 509.1 (580.4) | <.001 |
| P07225 | Vitamin K-dependent protein S | PROS1 | 81 | 8348.3 (2802.3) | 7095.1 (2604.1) | .02 |
| P55290 | Cadherin-13 | CDH13 | 77 | 5629.1 (7477.3) | 1672 (2380.7) | <.001 |
| P27348 | 14-3-3 protein theta | YWHAQ | 71 | 232.9 (214.4) | 328.2 (607) | .001 |
| Q9UGM5 | Fetuin-B | FETUB | 70 | 3058.5 (2968.6) | 2523 (3637.7) | .41 |
| Q00872 | Myosin-binding protein C, slow-type | MYBPC1 | 68 | 534 (318.2) | 1949.8 (2942.8) | <.001 |
| Q96RW7 | Hemicentin-1 | HMCN1 | 64 | 752.3 (1010.8) | 325.8 (394.3) | <.001 |
| P08246 | Neutrophil elastase | ELANE | 56 | 139.7 (223.9) | 766.3 (1561.7) | <.001 |
| P13667 | Protein disulfide-isomerase A4 | PDIA4 | 53 | 528.3 (1047.2) | 388.5 (743.2) | .001 |
| P18428 | Lipopolysaccharide-binding protein | LBP | 50 | 1475 (1208.9) | 2101.9 (2206.5) | <.001 |
| P21802 | Fibroblast growth factor receptor 2 | FGFR2 | 47 | 2438.8 (1974.7) | 749.8 (576.4) | <.001 |
| P50552 | Vasodilator-stimulated phosphoprotein | VASP | 44 | 85.7 (72.8) | 250.2 (304.5) | <.001 |
| A0A075B6I1 | Immunoglobulin lambda variable 4-60 | IGLV4-60 | 37 | 163.2 (476.425) | 231.3 (763.7) | .002 |
| P07203 | Glutathione peroxidase 1 | GPX1 | 33 | 138.7 (88) | 315.1 (299.3) | .001 |
| Q9NZD4 | Alpha-hemoglobin-stabilizing protein | AHSP | 32 | 193.3 (235.4) | 1344.5 (1473.8) | <.001 |
| A0A075B6K0 | Immunoglobulin lambda variable 3-16 | IGLV3-16 | 19 | 270.2 (488.3) | 497.6 (744.9) | <.001 |
| P53396 | ATP-citrate synthase | ACLY | 15 | 305 (565.9) | 1134.1 (1936.8) | <.001 |

### eFigures

#### eFigure 1. Neurological Assessment with EMS and NDI

The time course of spinal function assessed with EMS scale (**A**) and NDI (**B**), from baseline to 6 months postoperatively. There was no statistically significant difference in the change of all four scales between baseline and post-operatively follow-up at 3rd and 6th month.

**eFigure1.** Neurological Assessment with EMS and NDI

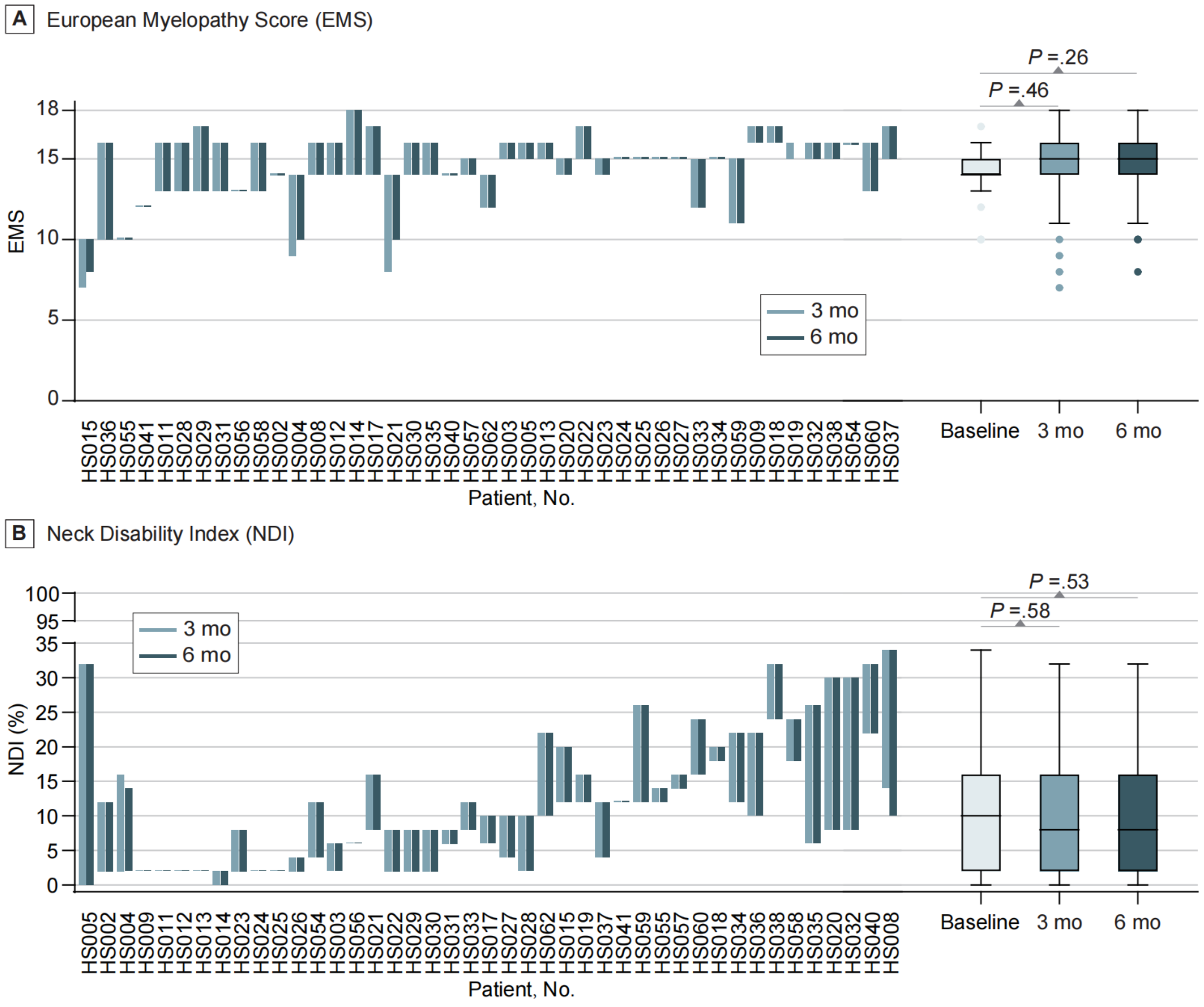

**eFigure 2. Biological Indications of Model Protein Variables**

**A**. Venn diagram. This diagram defined the protein whose variable importance score >0. The numbers of unique proteins in each group vs overlapped proteins were shown. Uniprot ID for all protein selected were list at the right panel. Proteins with variables importance score ≥ 50 were shown in bold font. **B**. Protein with importance scores ≥50 were enriched for interactions (connectivity *P* value of non-invasive group = 0.003, connectivity *P* value of invasive group = 0.40) as measured in the sum of unweighted degrees in the network. **C**. Gene ontology term analysis of genes corresponding to proteins with importance scores greater than 0 (adjusted *P* value <0.01).

**eFigure 2.** Biological Indications of Model Protein Variables

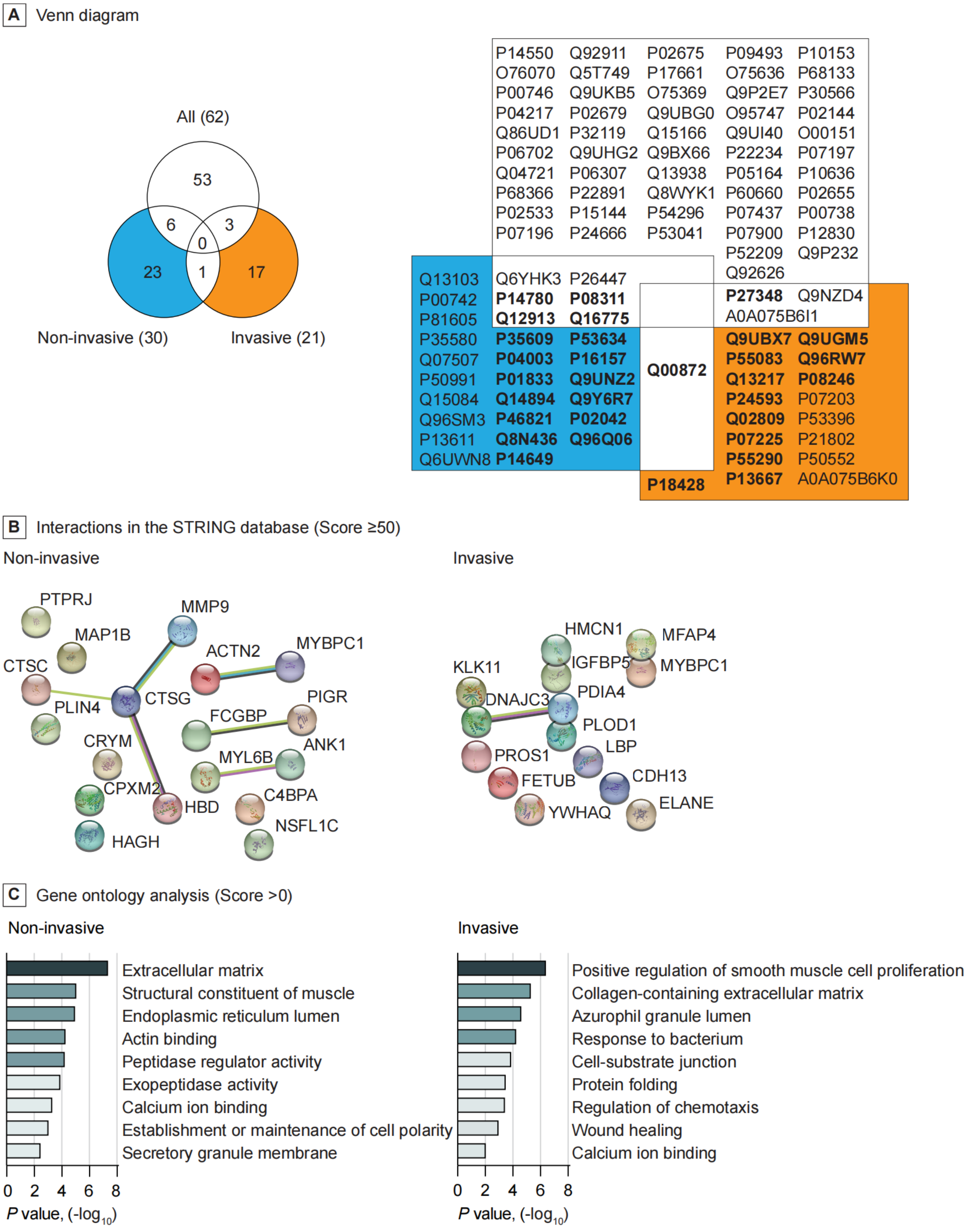

#### eMethods

##### Interaction Network analysis

Protein interaction was analyzed using STRING version 11 (STRING Consortium). Gene ontology terms were generated using the functional annotation tool Metascape (Metascape Team).
